# The extraordinarily long lead times of screen-detected lung cancer in never-smokers

**DOI:** 10.1101/2024.09.28.24314527

**Authors:** Wayne Gao, H Gilbert Welch

## Abstract

**Background:** The randomized trials of low-dose computed tomography (LDCT) screening enrolled individuals at high risk of lung cancer: heavy cigarette smokers. Nevertheless, opportunistic screening of never smokers has become commonplace in certain regions of East Asia. Lung cancer detection rates reported for this low risk group regularly exceed those reported in the randomized trials. We sought to determine the lead time – the length of time between screen detection and when symptoms would manifest (clinical presentation) – associated with LDCT detection rates reported in Taiwan.

**Objective:** To estimate the lead time for LDCT lung cancer detection in never smoking Taiwanese females, under the assumption that all detected cancers progress.

**Methods:** We analyzed data from the Taiwan Lung Cancer Screening in Never-Smoker Trial (TALENT), a prospective cohort of ≈12000 never smokers aged 55–75, and compared age-specific screen detection rates to lung cancer incidence rates in the pre-screening period. Lead time was estimated using Morrison’s approximation: one-half the ratio of screen detection to pre-screening incidence.

**Results:** Invasive lung cancer detection rates in never smoking females were 18 (age 70-74) to 37 (age 55-59) times higher than the respective age-specific lung cancer incidence rates prior to widespread screening. The estimated lead times for invasive lung cancers thus ranged from 9 (age 70-74) to 18.5 years (age 55-59). Had we included in situ lung cancer detection, the estimated lead time of screening would range from approximately 10 to 20 years.

**Conclusion:** If all detected cancers progress, the lead times for LDCT lung cancer detection in never-smokers would be 5 to 20 times longer than that reported in the randomized trials of heavy smokers (1 to 2 years). Such extended lead times are not only undesirable (*Why label and treat a patient for a cancer not destined to clinically present in the next 1 or 2 decades?*), but also a major source of overdiagnosis – providing plenty of time for a patient to die from other causes before developing a symptomatic cancer (Type 1 overdiagnosis). Furthermore, some screen-detected cancers may not progress and thus have an infinite lead time (Type 2 overdiagnosis). Regardless, it appears that substantial overdiagnosis has occurred in screening low-risk, never smoking individuals.

## Background

The goal of cancer screening is to reduce cancer mortality. To achieve this goal, screening must advance the time of cancer diagnosis from the point it would otherwise present clinically – in other words, it must introduce a lead time. Lead time is the length of time between screen detection and the time when symptoms would manifest (clinical presentation). Thus, lead time is a desirable aspect of screening – it is necessary, but not sufficient, for screening to lower mortality.

But lead times also create problems. The most familiar is lead time bias: the artifactual increase in survival time created simply by advancing the time of diagnosis even if the time of death is unchanged – in simple terms, “starting the clock earlier”. Lead time is the reason that survival statistics are always biased in the context of early detection and why mortality is the unbiased outcome metric. But long lead times also produce a less familiar problem: overdiagnosis. Long lead times provide plenty of time for a patient to die from other causes before developing a symptomatic cancer (leading to so-called Type 1 overdiagnosis).

For lung cancer screening, the lead times estimated in the major randomized trials are 2 years or less (NLST 1.3 to 2 years, NELSON 1.1 years).^1^ To minimize overdiagnosis, these trials enrolled patients with a high risk of lung cancer death (heavy smokers) and employed growth assessment protocols: instead of biopsying all pulmonary nodules immediately, small nodules were observed over time and only those that grew larger were biopsied. In East Asia, however, opportunistic screening of low-risk never smokers has become commonplace and growth assessment protocols are not employed. In this paper, we estimate the lead times associated with opportunistic screening in Taiwan.

## Methods

### Overview

Lead time is estimated as function of two variables: 1.) the incidence of clinical presenting, symptomatic lung cancer and 2.) the detection rate of lung cancer screening. High detection rates relative to lung cancer incidence imply longer lead times. The average lead time was estimated using the approach outlined by Morrison – which makes the simplifying assumption that all screen-detected cancers progress^2^. This assumption implies there are no screen-detected cancers with infinite lead times: that is, cancers that either do not progress or, in fact, regress (so-called Type 2 overdiagnosis). The lead time is approximated as one-half of the ratio between the screening detection rate and the lung cancer incidence rate^2^. For example, if the screen detection rate equals the lung cancer incidence, the mean lead time is six months; if the screen detection rate is double the annual incidence, the mean lead time is one year.

We estimate lead time for 4 screening age groups: age 55-59, age 60-64, age 65-69, and age 70-74.

### Data: Incidence of symptomatic lung cancer

To obtain age-specific data on the incidence of symptomatic lung cancer, we obtained lung cancer incidence data from the Taiwan National Cancer Registry for the year 2000^3^. This was prior to the promotion of widespread screening, ensuring that it reflects the incidence of clinically-detected cancer – cancers that present with symptoms.

To avoid the confounding effect of declining smoking prevalence in men, we restrict the analysis to women. Smoking prevalence has declined sharply among Taiwanese men since 1980 (from 60% to <25%),^4^ making 2000 incidence a considerable overestimate of the current male incidence of lung cancer destined to appear clinically. Smoking prevalence among Taiwanese women, however, has been stable since 1980 (less than 5%) making 2000 incidence an ideal estimate of the current female incidence of lung cancer destined to appear clinically.

### Data: Screen detection rate

We used data from the Taiwan Lung Cancer Screening in Never-Smoker Trial (TALENT), an observational cohort study involving approximately 12,000 never-smoking individuals aged 55-75 years. Lung cancer was diagnosed in 318 participants, corresponding to an overall detection rate of 2.6%, with 257 cases of invasive lung cancer (2.1%) and 61 cases of in situ cancers (0.5%)^5^. Here we examine the detection rate in the ≈4500 women <u>without</u> a family history of lung cancer and consider invasive cancers only (i.e. exclude in situ cancers).

The detection rate in females is inferred knowing that the overall detection rate in TALENT is the weighted average of the male and female detection rates (26% of TALENT participants without a family history were male and 74% were female) and knowing that detection rate in males was one-half that of females (OR = 0.5). For example, in the age group 55–59 years the detection rate for both sexes combined was 1.5%. To determine the detection rate in females (x) we solved the following equation:

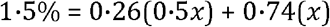

Thus, the inferred detection rate in females (x) was 1.73%. Similar calculations were repeated for females aged 60-64, 65-69, and 70-74 years.

## Results

As shown in the **Table**, the age-specific detection rates in TALENT were 18–37 times that of observed lung cancer incidence prior to widespread screening. Assuming all screen-detected cancers progress the estimated average lead times in TALENT are extraordinarily long: 9 years (age 70-74) to 18.5 years (age 55-59). Had we included in situ cancers, the detection rates would have increased (while the incidence of symptomatic cancer remained the same) and the estimated average lead times would have been approximately 10 to 20 years.

**Table:** Ratio of screen-detected lung cancer to pre-screening incidence in Taiwanese women.

| Age Group | Detection rate in TALENT* | Inferred detection rate in women† | Screen-detected lung cancer (per 100,000) | Pre-screening incidence in Taiwanese women (per 100,000) | Ratio of screen-detected to clinical presentation | Average Lead Time (in years) |
| --- | --- | --- | --- | --- | --- | --- |
| 55–59 years | 1.50% | 1.73% | 1730 | 47.1 | 37 | 18.5 |
| 60–64 years | 1.30% | 1.50% | 1500 | 63.4 | 24 | 12 |
| 65–69 years | 1.80% | 2.08% | 2080 | 92.7 | 22 | 11 |
| 70–74 years | 2.20% | 2.54% | 2540 | 139.9 | 18 | 9 |

\*No family history of lung cancer, both sexes.
†The detection rate in females is inferred knowing that the overall detection rate in TALENT is the weighted average of the male and female detection rates (26% of TALENT participants were male and 74% were female) and that detection in males is half that of females. For example, the equation for age 55–59 years is: $1.5\% = 0.26(0.5x) + 0.74(x)$ , where $x$ is the detection rate in females.
\*\*Incidence reported by the Taiwan National Cancer Registry for the year 2000.

## Discussion

The estimated lead times vary across common cancer screening tests: 1-2 years for lung cancer screening in heavy smokers, 1-4 years for breast cancer screening^6^ and 5-10 years for cervical^7^, colorectal^8^ and prostate cancer screening ^9^(also see Supplement). Thus, the estimated lead time of 10-20 years for lung cancer screening in never smoking Asian females is a remarkable outlier.

It is important to emphasize that these number are averages and obscure the underlying distribution of lead times. While it is impossible to measure the lead time of diagnosis in an individual, it is entirely possible that some never smoking Asian women are diagnosed with a lead time of 1 to 2 years. At the same time, that implies that others are diagnosed with a lead time of 20 to 40 years.

While cancer screening must introduce a lead time to effectively reduce mortality, extraordinarily long lead times are undesirable. Its hard to imagine why we would want to subject women to being labeled has having lung cancer – and being treated for lung cancer – 20 to 40 years before the cancer would otherwise become clinically evident. Furthermore, extraordinarily long lead times make Type 1 overdiagnosis extraordinarily likely: some women age 55 to 74 are certain to die of some other cause before their detected cancer has a chance to become clinically evident.

Finally, the estimation of lead time makes the simplifying assumption that anything labeled as being “cancer” will invariably progress. But some of these cancers – particularly the minimally invasive and in situ cancers – may not progress or, in fact, may regress (so-called Type 2 overdiagnosis). Regardless, the high lung cancer detection rates in screening low-risk, never smoking individuals in Taiwan suggests that substantial overdiagnosis has occurred.

## Data Availability

All data produced in the present work are contained in the manuscript

https://twcr.tw/?page_id=1843&lang=en

https://doi.org/10.1016/s2213-2600(23)00338-7

## Competing Interests

The authors declare no competing interests in relation to this study. Ethnics Statement: This study did not require ethics approval as it solely utilized publicly available secondary data from published research, focusing on lung cancer detection rate by age group and population cancer incidence from Taiwan cancer registry. The data were anonymized, with no individual identifiable information used, ensuring the privacy of individuals. As such, no informed consent was necessary for the analysis. The study adheres to all relevant ethical guidelines and respects the confidentiality of population-level data.

**Supplement: Estimated lead times of common cancer screening tests**

ChatGPT’s response to the question: *“What are the typical lead times of cancer screening tests?*”

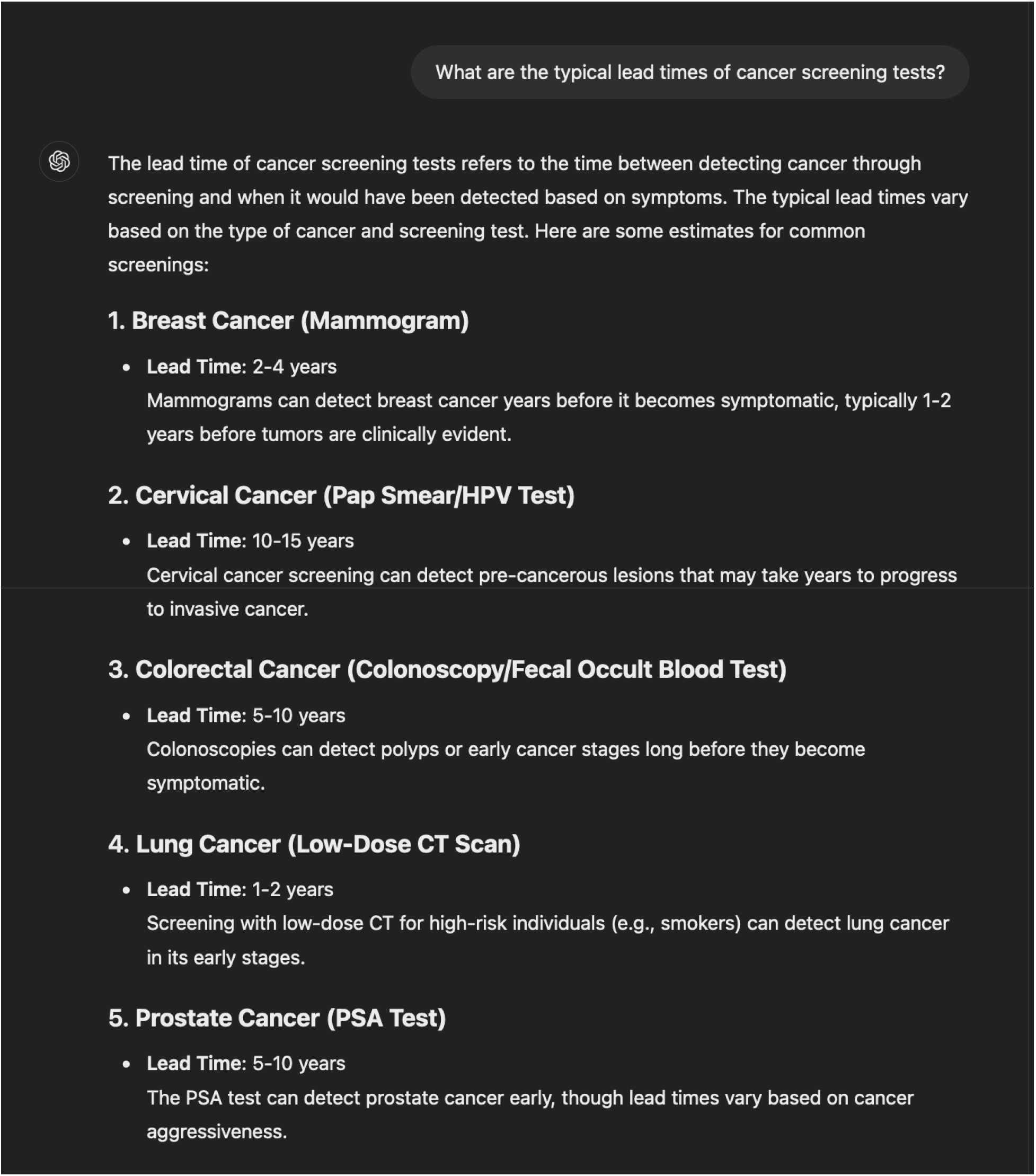

## References

1. Benbassat J. Duration of lead time in screening for lung cancer. BMC Pulmonary Medicine 2021; 21: 1–8.

2. Morrison AS. Screening in chronic disease; 1992.

3. Taiwan Cancer Registry Center. Incidence and mortality rates for top 10 cancers in Taiwan https://twcr.tw/?page_id=1855&lang=en. 2023. https://cris.hpa.gov.tw/pagepub/Home.aspx (accessed December 23 2023).

4. Health Promotion Administration Taiwan. Adult Smoking Behaviors Survey, (ASBS, Taiwan) 2021. https://www.hpa.gov.tw/Pages/Detail.aspx?nodeid=1718&pid=9913 (accessed Feb 13 2024).

5. Chang GC, Chiu CH, Yu CJ, et al. Low-dose CT screening among never-smokers with or without a family history of lung cancer in Taiwan: a prospective cohort study. Lancet Respir Med 2024; 12(2): 141–52.

6. Lee CI, Elmore JG. Beyond survival: a closer look at lead-time bias and disease-free intervals in mammography screening. JNCI: Journal of the National Cancer Institute 2023; 116(3): 343–4.

7. Holowaty P, Miller AB, Rohan T, To T. Natural history of dysplasia of the uterine cervix. J Natl Cancer Inst 1999; 91(3): 252–8.

8. Winawer SJ. Natural history of colorectal cancer. Am J Med 1999; 106(1a): 3S-6S; discussion 50S-1S.

9. Draisma G, Etzioni R, Tsodikov A, et al. Lead time and overdiagnosis in prostate-specific antigen screening: importance of methods and context. J Natl Cancer Inst 2009; 101(6): 374–83.

